# Health care utilization and the associated costs attributable to cardiovascular disease in Ireland: a cross-sectional study

**DOI:** 10.1101/2023.09.18.23295706

**Authors:** Danko Stamenic, Anthony P. Fitzgerald, Katarzyna A. Gajewska, Kate N. O’Neill, Margaret Bermingham, Jodi Cronin, Brenda M. Lynch, Sarah M. O’Brien, Sheena M. McHugh, Claire M. Buckley, Paul M. Kavanagh, Patricia M. Kearney, Linda M. O’Keeffe

## Abstract

**Background:** Cardiovascular diseases (CVD) are the leading cause of mortality and disability globally. An ongoing reform of the Irish healthcare system is underway with a focus on preventing avoidable CVD and lessening its burden to society. However, the high rates of healthcare service use attributable to CVD and the associated costs have not been adequately quantified in Ireland. We examined the difference in health service utilisation and costs for populations with and without CVD in Ireland for the period preceding the reform.

**Methods:** Secondary data analysis of the first wave (2009-2011) of The Irish Longitudinal Study on Ageing (TILDA), a nationally representative study of community-dwelling adults in Ireland aged 50+. CVD was defined as having a self-reported doctor’s diagnosis of myocardial infarction, angina, heart failure, stroke, atrial fibrillation or transient ischaemic attack. Participants self-reported the utilization of healthcare services over the 12 months preceding the interview. Negative binomial regression with average marginal effects (AME) was used to model the effect of CVD on healthcare service utilisation. We estimated the incremental number of general practitioner (GP) and outpatient department (OPD) visits, accident and emergency department (A&E) attendances and hospitalisations in population with CVD relative to population without CVD and calculated the associated costs. Analyses were adjusted for socio-demographic confounders and other chronic conditions. Using census 2022 data on the total number of people aged 50+ living in Ireland, we estimated the total incremental costs attributable to CVD at the population level.

**Results:** Among 8113 participants, the prevalence of CVD was 18.2% (95% confidence interval (CI): 17.3, 19.0). Participants with CVD reported higher utilization of all healthcare services. In adjusted models, having CVD was associated with incremental 1.19 (95% CI: 0.99, 1.39) GP and 0.79 (95% CI: 0.65, 0.93) OPD visits over the past year. There were twice as many incremental hospitalisations in males with CVD compared to females with CVD (AME: 0.20 (95% CI: 0.16, 0.23) for males vs AME: 0.10 (95% CI: 0.07, 0.14) for females), but no difference was observed with respect to the incremental use of other healthcare services by gender or age groups. The incremental cost of healthcare service use in this population relative to the population without CVD were an estimated €352.2 million (95% CI: €272.8, €431.7), 93% of which was due to use of secondary care services.

**Conclusion:** There are substantial use of healthcare services and costs associated with CVD in Ireland, with hospital admissions being the biggest contributor to costs. While a shift towards the management of uncomplicated CVD cases in primary care is currently being implemented in Ireland, continued efforts aimed at CVD primary prevention and management are required to contain healthcare service costs. Further research on gender-disparities in the use of healthcare services attributable to CVD is warranted.

## Introduction

Cardiovascular disease (CVD) is the leading cause of mortality and disability globally, with coronary heart disease (CHD) and stroke considered the two main contributors to disability in those aged over 50 years (1–3). It is estimated that over half a billion people currently live with CVD worldwide, and its prevalence has doubled in the past 30 years (3). In Ireland, CVD is the single greatest cause of death, currently accounting for one-third of all deaths and one in five premature deaths, with CHD and stroke accounting for the largest proportion of deaths due to CVD (4,5). Recent estimates from the fifth wave of the Healthy Ireland survey (2018-2019) indicate that the prevalence of CHD and stroke in people aged ≥ 45 years ranges from 4.2% to 14.5 % and 0.7% to 2.2% respectively, depending on the age group (6). Despite Ireland seeing a substantial decline in CVD mortality rates over the past 30 years (7,8), the overall disease burden has increased due to ageing populations and improved survival of those with CVD, with substantial implications for healthcare service use (4). Yet, the biggest part of the CVD burden is preventable through appropriate management of well-established risk factors, thus saving lives and easing the pressure on the healthcare system (9).

The role of GP and primary care in Ireland is crucial in the prevention of new cases of CVD and the management of established disease and has the potential to reduce the use of more costly acute care in a traditionally hospital-centred healthcare system (10). However, in Ireland, it is still required for abound 60% of the population to pay the full cost of GP care at point of use (11). This makes the financing model of Irish GP care highly unusual in the international context and often criticised on its equity and efficacy grounds. In Ireland, only those aged over 70, under 6 or with particular health needs that cause undue hardships are automatically eligible for a full Medical Card or a GP visit card which gives them access to free GP care, while the eligibility of others is assessed mainly based on income means test. There were only three people in ten with a Medical Card in 2018, with an additional one person in ten having a GP visit card (11). Another layer of complexity to the Irish healthcare system is added by the existence of private health insurance with the purpose of complementary, supplementary or duplicative cover, which both Medical/GP visit card holders and non-holders may avail of. In 2016, the Government introduced Sláintecare, a ten-year reform of the health and social care services in Ireland (11). The reform also considers a significant amendability to chronic disease prevention and healthcare (12), together with development of the Structured Chronic Disease Management Programme and the implementation of the Integrated Care Programme for the Prevention and Management of Chronic Disease (ICPCD) (10). Sláintecare aims to establish a universal single-tier healthcare service that ensures equitable access based on need rather than on the ability to pay and to move from the current hospital-centred system of healthcare provision to a primary and social care-focused system (12). While it is expected that the reform will also have an effect on how healthcare is delivered to those affected by CVD, the impact of CVD on the use of primary and secondary healthcare services and associated costs in the period before its introduction have not been assessed.

To date, studies in Ireland have predominantly examined healthcare service use in the context of specific CVD conditions in particular (e.g. stroke, coronary syndrome, angina, heart failure) (13,14) or in the context of the wider chronic disease burden (15,16) and without assessing the incremental use of healthcare services due to CVD (13–17). While some of these studies have evaluated the impact of CVD on specific aspects of healthcare service utilization, for example, hospitalisations due to ambulatory care sensitive conditions (15) or the variability in the secondary care prevention in outpatients with coronary syndrome (14), they did not examine the costs associated with the increased health service utilisation for this cohort (13,14,16,17). In contrast, the prevalence of diabetes in Ireland is well documented (6,18–20) and the burden of diabetes and its complications has been extensively studied both in terms of higher consumption of healthcare services and the associated costs (15,19,21–23).

Using a nationally representative sample of community-dwelling adults in Ireland aged 50+, the aim of this study is to estimate the incremental healthcare service use attributable to CVD and the associated costs prior to the introduction and the implementation of Sláintecare and ICPCD to the Irish healthcare system.

## Methods

This study is reported using the strengthening of the reporting of observational studies in epidemiology (STROBE) guidelines for reporting observational studies (24).

### Data

The Irish Longitudinal Study on Ageing (TILDA) is a large-scale nationally representative prospective study of the community-dwelling population of Ireland aged 50 years and above (25,26). The first wave of TILDA recruited a stratified clustered sample of 8,175 individuals using the RAMSAM system which relies on the Irish Geodictionary, an up-to-date and comprehensive list of residential addresses in Ireland (26,27). Briefly, all postal addresses in Ireland were assigned to one of 3,155 geographic clusters, stratified by geography and socio-economic group and a sample of 640 clusters was selected with a probability that is proportional to the number of individuals aged 50+ years in each cluster. Forty households were then selected from each cluster to allow for 25,600 addresses to achieve a sample size of 8,000, and each address was visited by an interviewer to establish eligibility criteria. All the individuals aged 50+ living at the selected addresses at the time of the visit together with their partners (of any age) were invited to take part in the study. Out of 10,128 addresses that were considered eligible for inclusion based on the presence of an individual aged 50+ years, successful interviews were obtained from 6,279 addresses (62% response rate) and 8,175 participants aged 50+ years were recruited. Ethical approval was obtained from the Trinity College Dublin Research Ethics Committee. Data collection took place between October 2009 and November 2011. All individuals who agreed to participate in the study were visited by a trained interviewer and underwent a Computer-Assisted Personal Interview (CAPI) in their home with questions relating to sociodemographic information, general health and well-being as well as self-reported doctor diagnosis of chronic conditions and healthcare utilization.

Participants were given a card with a list of heart conditions and asked whether a doctor had ever told them that they had any of the conditions on the card. We considered a participant as having CVD if they reported a doctor’s diagnosis of any of the following conditions: heart attack (myocardial infarction), angina, heart failure (congestive cardiac failure), stroke (cerebrovascular accident), atrial fibrillation (abnormal heart rhythm) or ministroke (transient ischaemic attack). Participants reported the number of visits to the GP and outpatient department (OPD), the number of attendances to accident and emergency department (A&E) and the number of hospital admissions in the 12 months preceding the interview.

Other variables of interest included age, gender, marital status (married or living with partner/never married/separated or divorced/widowed), highest education level attainment (none or primary/secondary/tertiary), household location (Dublin/another town or city/rural), self-reported health status (excellent/very good/good/fair/poor) (28), healthcare cover (Medical or GP visit card/private health insurance only/dual cover/none) and self-reported doctor diagnosis of other chronic conditions deemed not to be associated with CVD. Participants who reported having a medical or GP visit card were considered as having public health insurance. Between 2009 and 2011 (e.g. period during which data collection took place), the eligibility for a medical or GP visit card in Ireland was primarily based on an income means test, with the exception of those aged 70+ years (29,30).

### Analysis

We examined healthcare service utilization of participants with and without CVD. The differences in the mean number of GP and OPD visits, attendances at A&E and hospital admissions were compared using Wolcoxon rank sum test and the differences in the proportion of people attending each type of healthcare service at least once using Pearson’s chi-square test. Ordinary least squares, Poisson, negative binomial, zero-inflated Poisson and zero-inflated negative binomial regressions were used to model the association between CVD and the frequency of healthcare service use. The choice of the model was based on the comparison of Akaike Information Criterion (AIC) and the inspection of model predicted vs. observed probabilities. Negative binomial regression was selected for all four studied outcomes based on the lowest AIC values and the best fit to the data (Supplementary Figure 1 and Supplementary Table 1) and average marginal effects (AME) were calculated to provide an estimate of the incremental GP and OPD visits, A&E attendances and hospital admissions that were attributable to CVD.

The marginal effect of CVD is the change in the expected number of outcomes (that is number of GP and OPD visits, A&E attendances and hospital admissions) due to CVD for each individual in the dataset based on his/her observed set of values for variables included in the model. The AME averages the marginal effects over all subjects and represents the average number of events due to CVD (31).

### Covariates

The inclusion of appropriate variables in the multivariable regression models was informed by the Anderson framework for the societal and individual determinants of healthcare utilization with the aim of identifying an independent effect of CVD on healthcare service utilization (32). Accordingly, multivariable models were adjusted for age, gender, marital status (predisposing factors), location, highest education level attainment, health care cover (enabling factors) and other chronic conditions including diabetes, asthma, chronic obstructive pulmonary disease, osteoporosis, arthritis, cancer and ulcers (need factors), while any potential mediators of the association between CVD and healthcare service use were omitted. We included interaction terms between CVD and age and between CVD and gender to allow the effect of CVD on healthcare service utilization to vary by age and gender. Sampling weights were applied to all estimates in order to adjust for differential non-response and to reduce the potential for selection or participation bias (26).

### Cost data

Previously reported average unit costs of €50 for a GP visit, €160 for an OPD visit, €5,030 for hospital inpatient admission and €183 for A&E attendance for Ireland were used (33,34). We inflated these to represent the costs in Euro for April 2023 using the Consumer Price Index Inflation Calculator for Ireland (35). The corresponding inflated costs were €58.3, €191, €5,991 and €225 for a GP visit, OPD visit, A&E attendance and hospital admission, respectively. The average cost per person of the incremental healthcare service use attributable to CVD was calculated by applying AME to the corresponding average unit costs for the relevant healthcare service type. We also estimated the AME and average costs per person for males, females and age sub-groups (50-59, 60-69 and 70+ years old) in the sample. These costs were then extrapolated to the total Irish population with CVD by applying the prevalence estimate of CVD in the sample (e.g. age and gender-specific, where necessary) to the most recent Irish census data (2022) to obtain costs attributable to the incremental healthcare service use attributable to CVD at population level.

All analysis was carried out in R version 4.1.2 (36). We carried out the complete case analysis as complete data was available from 8,113 participants (99.2%) (Figure 1).

**Figure 1:**
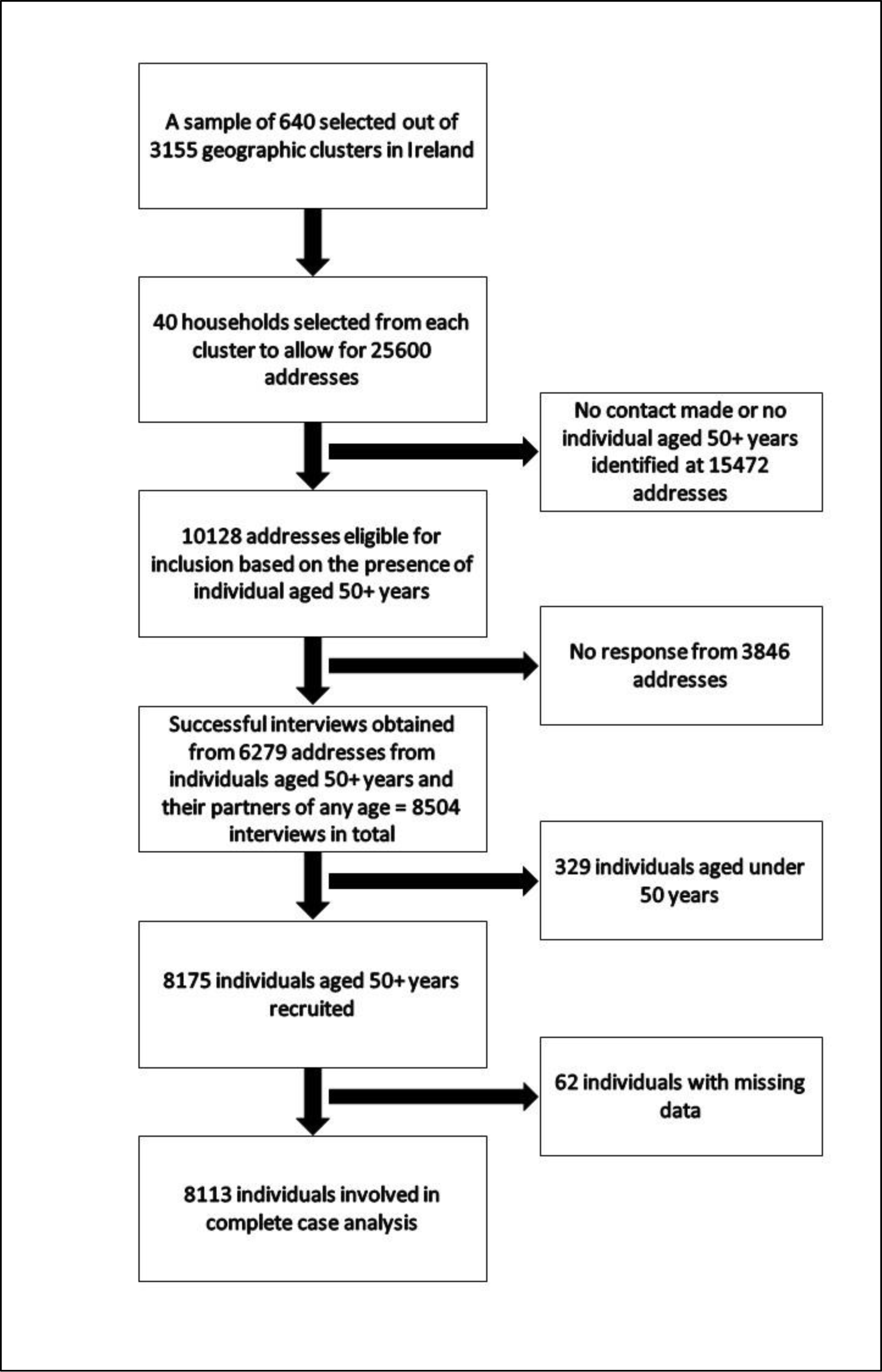
A flow diagram of participant inclusion in the TILDA study

## Results

The mean age of the participants was 63.5 years and 54.2% were female. The prevalence of CVD was 18.2% (95% confidence intervals (CI): 17.3, 19.0), and 5.6% (95% CI: 5.1, 6.1) had more than one cardiovascular condition. Characteristics of participants by their CVD status are shown in Table 1. Compared to the participants without CVD, those with CVD were on average older and were more likely to be male, widowed, less educated, have another chronic condition and reside in urban areas. They were less likely to report good health and have private health insurance only.

**Table 1:**
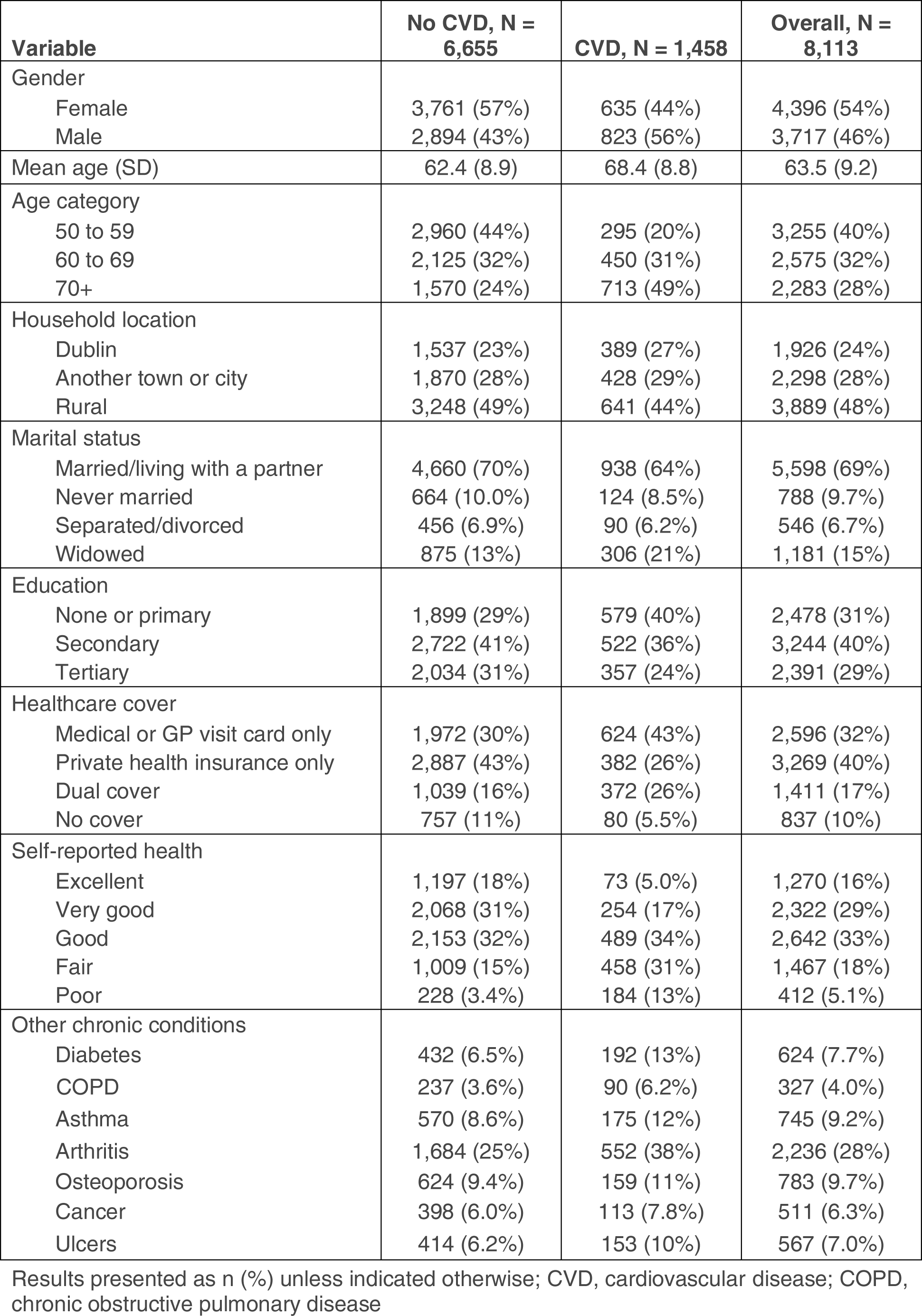
Characteristics of the study population by CVD diagnosis.

A greater proportion of individuals with CVD reported seeking healthcare services than those without CVD (Table 2). Specifically, a greater percentage of participants with CVD reported visiting a GP (95% (95% CI: 94%, 96%) vs 86% (95% CI: 85%, 87%)) and OPD (60% (95% CI: 58%, 63%) vs 37% (95% CI: 36%, 38%)), attending A&E (24% (95% CI: 22%, 26%) vs 13% (95% CI: 12%, 14%)) or having a hospital admission (25% (95% CI: 23%, 27%) vs 10% (95% CI: 9.6%, 11%)) over the 12 months preceding the interview.

**Table 2:** Use of healthcare services over the past year by CVD diagnosis.

| Healthcare service | No CVD, N =<br>6,655 | CVD, N = 1,458 | Overall, N =<br>8,113 |
| --- | --- | --- | --- |
| <u>GP visits</u> |  |  |  |
| Mean (SD) number | 3.4 (3.8) | 5.7 (5.0) | 3.9 (4.1)<sup>***</sup> |
| Attended | 5,703 (86%) | 1,391 (95%) | 7,094 (87%)<sup>***</sup> |
| <u>A&E attendance</u> |  |  |  |
| Mean (SD) number | 0.2 (0.6) | 0.4 (0.9) | 0.2 (0.7)<sup>***</sup> |
| Attended | 871 (13%) | 348 (24%) | 1,219 (15%)<sup>***</sup> |
| <u>OPD visits:</u> |  |  |  |
| Mean (SD) number | 1.0 (2.0) | 2.1 (2.8) | 1.2 (2.2)<sup>***</sup> |
| Attended | 2,446 (37%) | 880 (60%) | 3,326 (41%)<sup>***</sup> |
| <u>Hospital admissions:</u> |  |  |  |
| Mean (SD) number | 0.2 (0.6) | 0.4 (0.9) | 0.2 (0.6)<sup>***</sup> |
| Admitted | 686 (10%) | 362 (25%) | 1,048 (13%)<sup>***</sup> |

Results presented as n (%) unless indicated otherwise; CVD, cardiovascular disease; GP, general practitioner; A&E, accident and emergency department; OPD , outpatient department; <sup>\*\*\*</sup> indicate significance at level $p < 0.001$ .

AME from unadjusted and adjusted negative binomial regression models are presented in Table 3. Having CVD was associated with higher incremental healthcare service use across all studied services. In the adjusted model, having CVD was associated with on average 1.19 (95% CI: 0.99, 1.39) incremental GP visits and 0.79 (95% CI: 0.65, 0.93) incremental OPD visits over the past 12 months. Having CVD was also associated with 0.14 (95% CI: 0.10, 0.18) incremental A&E attendances and 0.15 (95% CI: 0.11, 0.18) incremental hospital admissions over the past year.

**Table 3:**
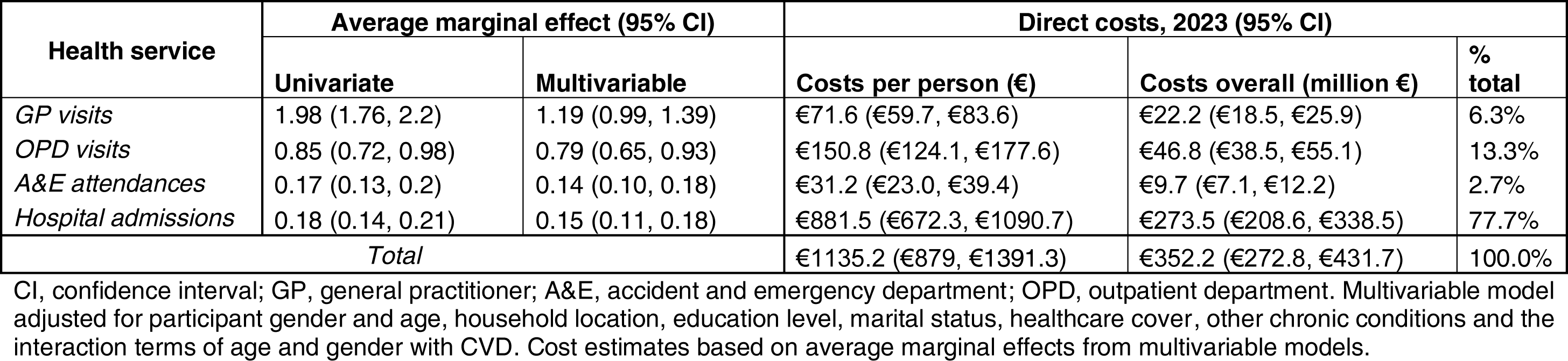
Incremental healthcare service use and costs attributable to CVD. Outputs from univariate and multivariable analysis.

Table 3 also provides estimates of the mean per capita costs and population-level costs attributable to incremental healthcare services use associated with CVD in Ireland. These costs are also presented according to healthcare service type. There was an estimated average cost of €1,135 (95% CI: €879, €1,391) per person associated with the incremental healthcare service use attributable to CVD. Applying the prevalence of CVD in the sample to the total Irish population aged 50+ (CSO estimate for 2022: 1,696,153) it was estimated that 310,305 adults aged 50+ in Ireland live with CVD. The cost of the corresponding incremental healthcare service use associated with CVD in this population was €352 million (95% CI: €273 million, €432 million) per annum. Hospital admissions accounted for nearly 80% of this cost (estimate: €274 million; 95% CI: €209 million, €339 million), with OPD visits accounting for an additional 13% (estimate: €46.8 million; 95% CI: €38.5 million, €55.1 million).

Figure 2 displays the incremental healthcare service use associated with CVD and the corresponding costs separately for males and females. While there was no difference between males and females in the incremental GP visits, OPD visits and A&E attendances associated with CVD, males had higher incremental hospital admissions attributable to CVD compared to females (AME: 0.20 (95% CI: 0.16, 0.23) in males vs AME: 0.10 (95% CI: 0.07, 0.14) in females). The total per capita cost of the incremental healthcare service use attributable to CVD was 72% higher in males than females (€1,470 (95% CI: €1,214, €1,726) vs €852 (95% CI: €596, €1,108)), and the difference was mainly driven by higher cost of incremental hospitalizations in males with CVD compared with females with CVD (€1,183 (95% CI: €974, €1,392) in males vs €627 (95% CI: €418, €826) in females). When calculated at population level, taking into account the gender-specific prevalence of CVD in the sample (estimate: 21.0% (95% CI:19.7%, 22.4%) in males and 15.6 % (95% CI: 14.4%, 16.7%) in females), the costs associated with the incremental healthcare service use attributable to CVD were 119% higher in males than in females (€254 million (95% CI: €210 million, €298 million) vs €116 million (95% CI: €81, €151 million).

**Figure 2:**
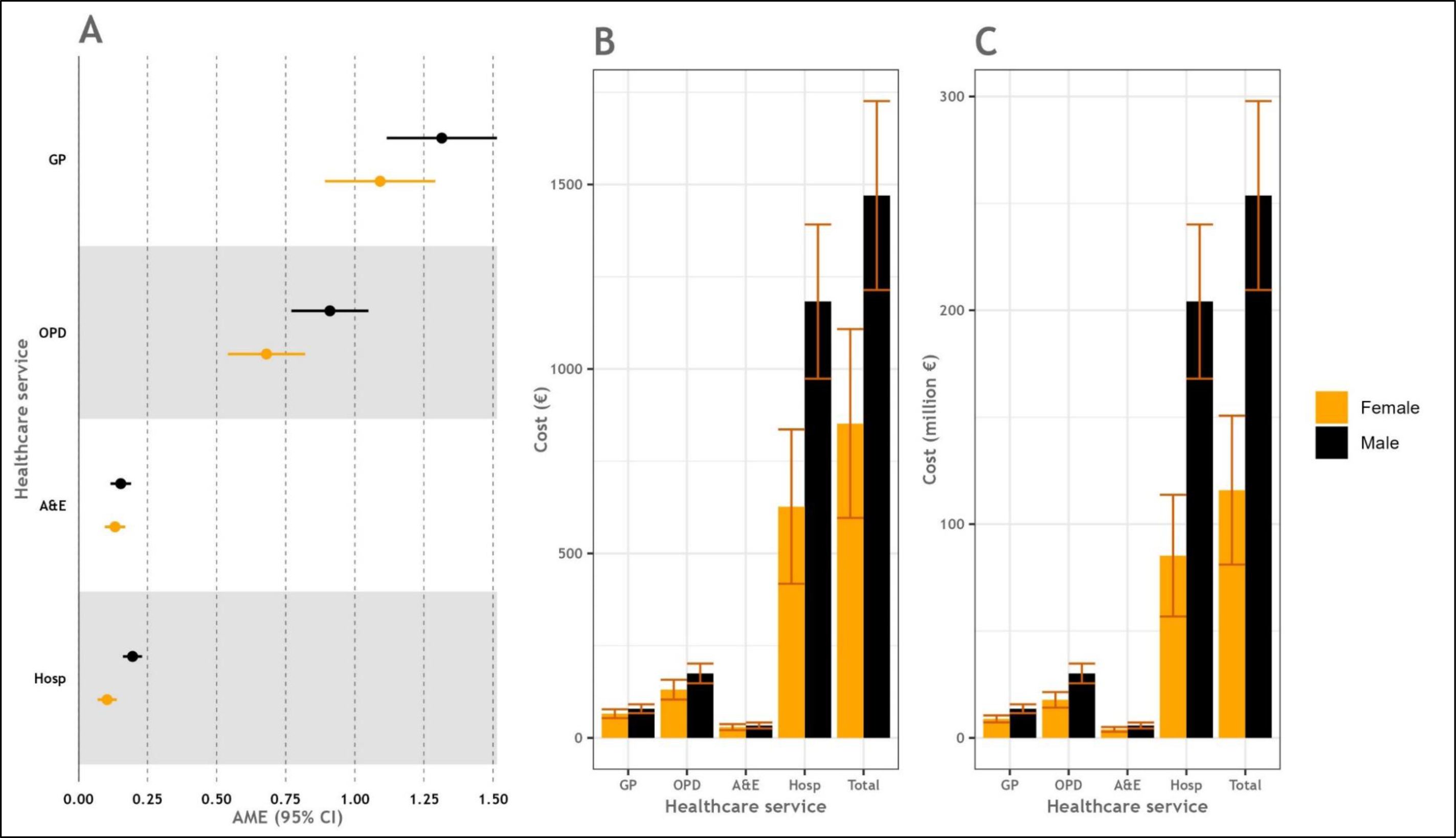
Incremental use of healthcare services attributable to CVD and associated costs stratified by gender. (A) Average marginal effects of CVD on healthcare services use, (B) Cost per person of incremental healthcare services use attributable to CVD, (C) population-level costs of incremental healthcare services use attributable to CVD. Estimates are given with 95% CI. CVD, cardiovascular disease; AME, average marginal effect; CI, confidence intervals; GP, general practitioner; OPD, outpatient department; A&E, accident & emergency department; Hosp, hospital admissions.

There were no differences observed in the incremental healthcare service use attributable to CVD between those aged 50-59, 60-69 and 70+ years (Figure 3). However, an increasing trend in cost at the population level was observed when moving from the youngest to the oldest group both in total costs (€62.3 million (95% CI: €44.2 million, €80.4 million) vs €103 million (95% CI: €79.1 million, €127 million) vs €208 million (95% CI: €132 million, €283 million) for age groups 50-59, 60-69 and 70+ years) and across each studied healthcare service separately (Figure 3). This was however mainly driven by a higher prevalence of CVD in older age groups (9.3% (95% CI: 8.3%, 10.4%) vs 17.6% (95% CI: 16.1%, 19.1%) vs 31.3% (95% CI: 29.3%, 33.3%) for age groups 50-59, 60-69 and 70+ years).

**Figure 3:**
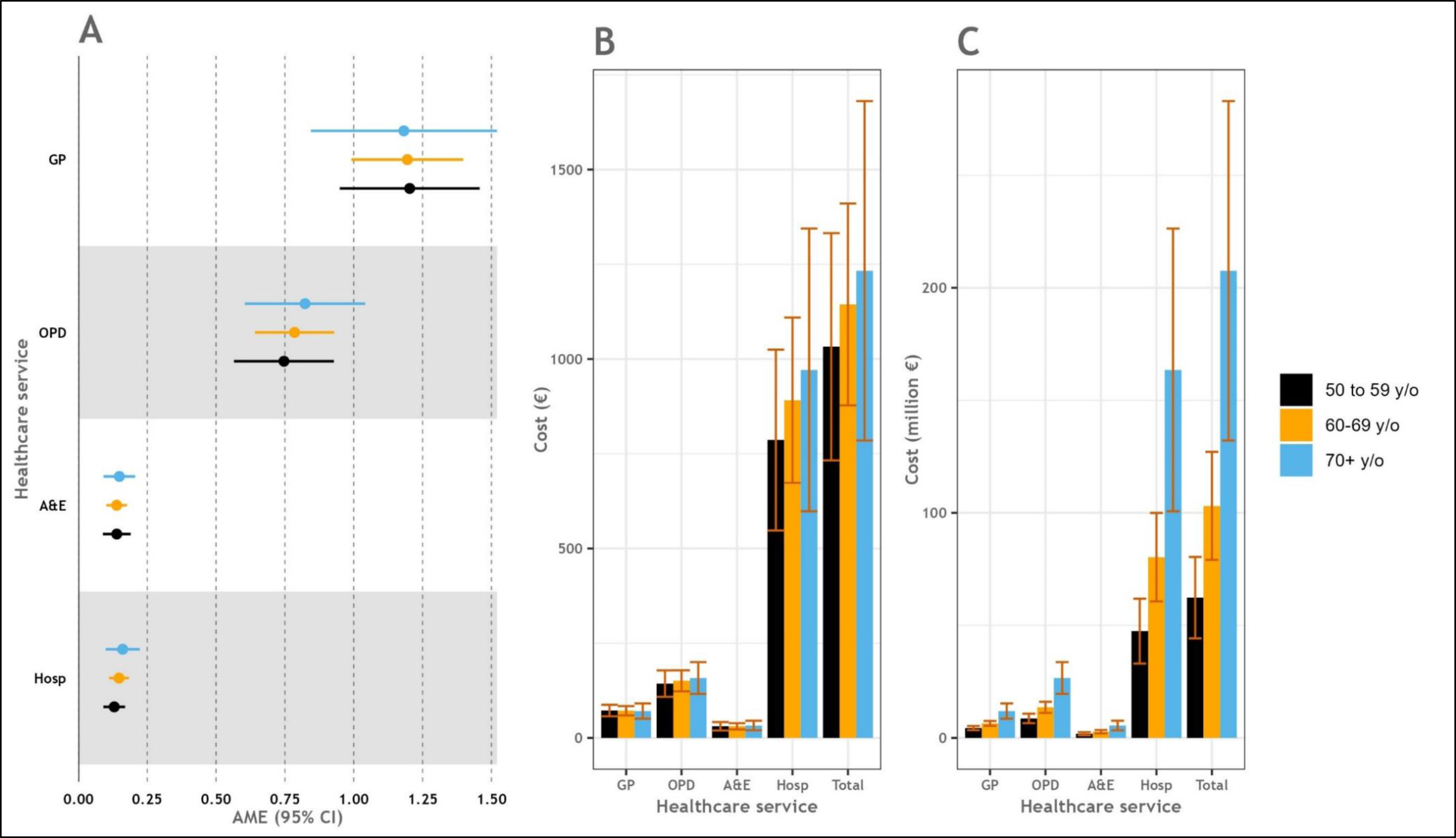
Incremental use of healthcare services attributable to CVD and associated costs stratified by age group. (A) Average marginal effects of CVD on healthcare services use, (B) Cost per person of incremental healthcare services use attributable to CVD, (C) population-level costs of incremental healthcare services use attributable to CVD. Estimates are given with 95% CI. CVD, cardiovascular disease; AME, average marginal effect; CI, confidence intervals; GP, general practitioner; OPD, outpatient department; A&E, accident & emergency department; Hosp, hospital admissions

## Discussion

In this large nationally representative sample of older adults in Ireland from 2009-2011, having CVD was associated with higher incremental healthcare service utilization across all services examined, incurring an estimated total marginal cost of €352 million at the population level and an estimated marginal cost of €1,135 per person per annum at the individual level. The main contributor to costs were hospital admissions which accounted for more than three-quarters of the costs. While there is a paucity of studies on the cost of CVD in Ireland, this finding is in line with the research on healthcare expenditure due to CVD reported internationally, whereby hospital inpatient costs are the main contributor to total healthcare costs (37–40). In addition, we observed increased hospital admissions associated with CVD in males compared to females and individual and population-level costs attributable to CVD were higher in males due to greater healthcare service utilization and higher prevalence of CVD in this population. An increasing trend was also observed in costs of the healthcare services associated with CVD at population level when moving from the youngest to the oldest age group, but this was driven by the increase in prevalence of CVD in older ages.

To the best of our knowledge, this study represents the first examination of the incremental healthcare service use attributable to CVD in Ireland, as well as the associated costs. Our analysis considers both primary and secondary healthcare costs, thus providing a comprehensive understanding of the financial implications of CVD on the Irish healthcare system. Direct comparisons with previous studies in Ireland are challenging due to differences in methodologies. Previous studies examined selected CVD endpoints (13,14,41) or wider chronic disease burden (15,16), focusing on specific aspects of secondary care only (14,15) or not calculating the incremental health service use attributable to CVD and associated costs (13,14,16,17). However, our results are broadly comparable with an analysis of 15,673 individuals from the 2010 Irish Quarterly National Household Survey, which at a similar time point to our study showed higher use of GP services and inpatient hospital care in individuals with CVD and CVD-related or non-CVD-related comorbidity over the 12 months preceding the survey (17). In another study analysing the same cohort as this study (i.e. the TILDA cohort) the impact of stroke, cognitive decline and post-stroke cognitive impairment on healthcare utilization was examined (13). After adjusting for demographic and clinical covariates, participants with stroke were more likely to visit a GP or OPD in the 12 months preceding their interview than participants without stroke, but there was no association between stroke and A&E visits or number of nights spent in hospitals (13). While this is partially in line with the findings from our study on the use of incremental GP and ODP services due to CVD, we also found higher attendances to A&E and higher hospital admissions attributable to CVD. While we cannot completely exclude the possibility that this discrepancy is due to the difference in the healthcare utilization between the participants with stroke and the participants with other CVD, there was a relatively small number of participants with stroke in the 1^st^ wave of TILDA (n=133, 1.6%) thus potentially limiting the power of certain analyses in the study by Jeffares *et al.*, as acknowledged by the authors (13). Our findings further build on these studies by providing estimates of the incremental healthcare service use attributable to CVD with the associated costs overall as well as by gender and age group.

The number of hospital admissions attributable to CVD was twice as high in males than in females, with the cost per person associated with the incremental healthcare services use being 72% higher in males than in females. Significant gender disparities have been reported with respect to the presentation, prevention and management of CVD internationally, although with conflicting results (42–45). An Australian study that reviewed the records of >50,000 patients from 60 primary healthcare services for the treatment of CVD risk found that, after adjustment for demographic and clinical characteristics and relative to males, females had lower odds of having sufficient risk factors measured for CVD risk assessment (OR:0.88; 95%CI: 0.81, 0.96), with heterogeneity in the gender-specific prescription of guideline-recommended medications across different age groups (42). Another study that involved 503 cardiologists in simulated decision-making for suspected coronary artery disease revealed that cardiologists have varying degrees of implicit gender bias for the characteristics of strengths and risk taking in patients with an immediate likelihood of coronary heart disease (43). In a large retrospective cohort of US adults, females were less likely to fill a guideline-recommended prescription for high-intensity statins following hospitalization for myocardial infarction compared to males (44). In another US study with over 20,000 participants of the Medical Expenditure Panel Survey, compared to their male counterparts, female adults with atherosclerotic CVD were less likely to report aspirin or statin use and more likely to utilize the emergency department two or more times per survey year (45). They were also more likely to experience lower healthcare satisfaction, poor perception of health status, poor patient-provider communication and lower health-related quality of life (45). To the best of our knowledge, the findings of our study are among the first that suggest a gender disparity in the use of healthcare services attributable to CVD Ireland. Further research is required to confirm these findings.

The higher use of GP services among individuals with CVD observed in our study suggests that a significant proportion of the burden of CVD is managed through primary healthcare services. This is in line with the chronic nature of CVD whereby the majority of its burden is expected to be managed in primary care, with exacerbations often requiring acute care (10). It is however interesting to note that just over 93% of costs associated with the incremental healthcare service use attributable to CVD at the population level relates to secondary healthcare services. Sláintecare, a 10-year programme, is a proposed reform of the Irish healthcare system introduced in 2016. The aim of Sláintecare is to establish a universal single-tier healthcare service that ensures equitable access based on need rather than on the ability to pay and to move from the current hospital-centred system of healthcare provision to a primary and social care-focused system (12). In terms of management of CVD, this translates to managing all uncomplicated cases of already established disease in the primary healthcare setting where GPs act as gatekeepers to the secondary system thus reducing the number of unnecessary hospitalisations. While this shift is currently ongoing, continued efforts aimed at CVD primary prevention are required for the conditions that cannot be managed in primary care (e.g. heart attack) coupled with health promotion to further tackle the CVD burden on the Irish healthcare system (46). Preliminary results from the Structured Chronic Disease Management Programme in Ireland are indicative of some improving trends over time in both self-reported and non-self-reported risk factors for CVD in this context (47). The national implementation of the Integrated Model of Care for the Prevention and Management of Chronic Disease also emphasizes secondary prevention, self-management support and the management of more complex CVD and multimorbidity in the community as important factors in reducing the burden of CVD in Ireland (10). The implementation of Sláintecare together with the reorganization of the Irish healthcare system should be evidence-informed and its performance assessed relative to the period prior to its introduction. As a period of considerable reform of the Irish health system is ongoing, our findings are of direct relevance in that they provide information on healthcare utilization among older adults with CVD in a period before the introduction of Sláintecare and particularly the ICPCD and can serve as a baseline for future evaluations of healthcare services use for this cohort. Further analyses should establish the health service use attributable to CVD in the more recent years and contrast these to our findings from 2009-2011.

It is important to consider the potential limitations of the current study when interpreting the findings. We cannot exclude the possibility of reverse causality due to the cross-sectional nature of the study, that is the participants who attended the GP were more likely to be diagnosed with CVD. Nevertheless, 87 % of the cohort reported visiting their GP at least once over the year preceding the interview thus reducing the potential for reverse casualty. We also acknowledge that misclassification bias may have been introduced by individuals reporting their doctor’s diagnosis of CVD. It has however been shown that self-report is a suitable measure for estimating prevalence of CVD compared to electronic medical records, with a substantial agreement between the two measures for stroke and myocardial infarction and a moderate agreement for heart failure (48). In addition, healthcare services utilization was also self-reported in the current study introducing the potential for measurement bias. Self-report is however often used in health services research and while evidence suggests a good validity of self-reporting for GP and OPD visits (49,50), the number of emergency department visits was found to be slightly over-reported in one of the studies using data from TILDA (49). We anticipate that this would have unlikely impacted our main findings as there were on average only 0.14 incremental A&E visits associated with CVD accounting for less than 3% of the total costs attributable to CVD at a population level. Furthermore, while we aimed to explore the cost of the incremental healthcare service utilisation attributable to CVD, we acknowledge that the total cost of CVD in Ireland is likely much higher. Other categories of healthcare costs relating to CVD such as medication have been found to contribute to this. For instance, medication expenditure was shown to account for between 11% and 52% of direct healthcare costs attributable to CVD across Europe which came mainly secondary to hospital inpatient costs but greater than the costs of GP, OPD and emergency department visits (40). Lastly, data collection for the first wave of TILDA used in our study took place between 2009 and 2011. However, the results on the incremental healthcare service utilisation attributable to CVD are still of relevance as they refer to the period before the introduction of the Chronic Disease Management Programme in Ireland and can serve as a baseline assessment for the programme. Our study also has several strengths including the comparison of CVD to no CVD to assess the incremental use of healthcare services attributable to CVD and the inclusion of both primary and secondary healthcare services in the analyses. Our results are also representative of the Irish general population over 50 years and using census 2022 data we have provided the total incremental costs attributable to CVD at the population level. Another strength of our study is reporting on AME. These are easily understandable and straightforward to interpret contrary to incidence rate ratios that are the traditionally reported effect size from count data models and that only provide a relative measure of the relationship between exposure and outcome, without giving the true sense of the magnitude of effect (51,52). Lastly, in order to adjust for differential non-response, minimize the potential for selection bias and improve the representativeness of our findings, survey weights were applied to all the estimates.

## Conclusion

Our findings suggest that while most CVD burden in Ireland is managed through primary care more than 93% of the incremental costs attributable to CVD are generated through the use of secondary care services. While a shift towards the management of uncomplicated CVD cases in primary care is currently being implemented in Ireland, continued efforts aimed at CVD primary prevention and management are required to reduce health service costs attributable to CVD. Further research with regard to gender disparity in the use of healthcare services due to CVD is required.

## Data Availability

All data produced in the present study are available upon reasonable request to the authors.

## Source of support

This work was supported by a research grant from the Irish Health Research Board (reference: SDAP-2019-030). The funders had no role in the study design, data collection and analysis, decision to publish or preparation of the manuscript.

## Author Contributions

D.S. had a primary responsibility for writing the manuscript. D.S. performed the analysis. D.S., A.P.F., P.M.K. and L.M.O.K contributed to the interpretation of data and analyses. All authors made a substantial contribution to the conception or design of the work. All authors contributed to the final manuscript, provided its critical revision and gave their approval for the final version to be published.

## Conflict of Interest

No competing interests exist.

## Supplementary figures

**Table S1:** Model diagnostics - Akaike Information Criteria for different structural models.

| **Model** | **Ordinary Least**<br>**Squares** | **Poisson** | **Zero Inflated**<br>**Poisson** | **Negative**<br>**Binomial** | **Zero Inflated**<br>**Negative**<br>**Binomial** |
| --- | --- | --- | --- | --- | --- |
| General Practitioner visits | 46370.33 | 51318.02 | 48866.65 | 40045.3 | 40047.58 |
| Accident & Emergency Department visits | 16945.26 | 10405.19 | 9454.011 | 9207.58 | 9209.581 |
| Outpatient Department visits | 36021.88 | 32725.25 | 25885.44 | 23549.9 | 23551.56 |
| Hospital overnight admissions | 15872.74 | 9473.855 | 8504.034 | 8299.041 | 9477.854 |

**Figure S1:**
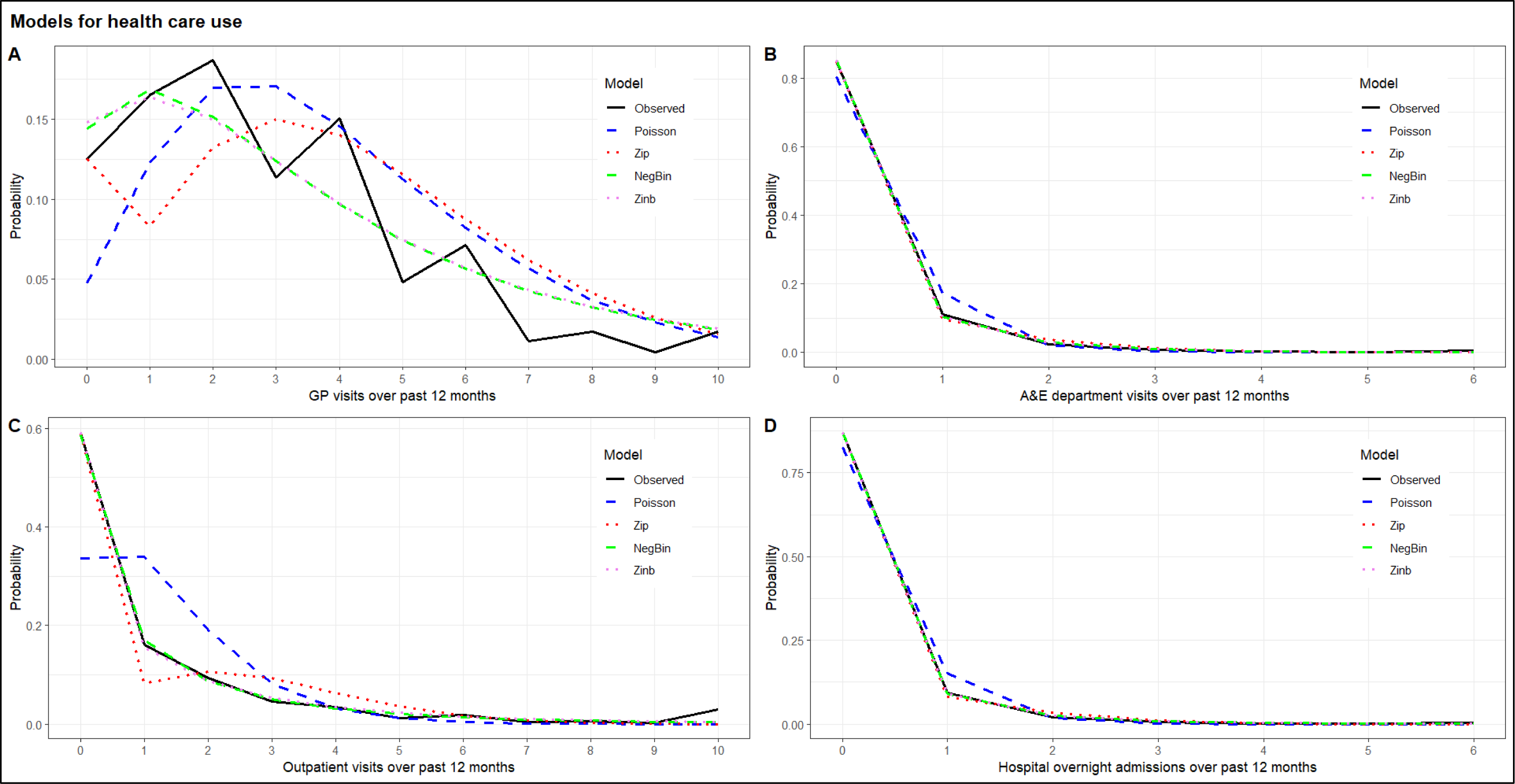
Observed and predicted probabilities for (A) number of GP visits, (B) number of accident & emergency department attendances, (C) number of outpatient department visits and (D) number of hospital overnight admissions. Zip, zero-inflated Poisson; NegBin, negative binomial; Zinb, zero-inflated negative binomial

## Notes

### Competing Interest Statement

The authors have declared no competing interest.

### Author Declarations

The study used only openly available data that ware originally located at: https://www.ucd.ie/issda/data/tilda/

